# Liver fibrosis assessed using non-invasive markers and genetic polymorphisms (*PNPLA3* and *TM6SF2*) predisposing to liver fibrosis, is associated with hospitalization or death from heart failure: a prospective UK Biobank study

**DOI:** 10.1101/2023.08.23.23294514

**Authors:** Theresa J Hydes, Oliver J Kennedy, Kate Glyn-Owen, Ryan Buchanan, Julie Parkes, Daniel J Cuthbertson, Paul Roderick, Christopher D Byrne

**Affiliations:** Department of Cardiovascular and Metabolic Medicine, Institute of Life Course and Medical Sciences, Faculty of Health & Life Sciences, University of Liverpool; University Hospital Aintree, Liverpool University Hospitals NHS Foundation Trust, Liverpool; Liverpool Centre for Cardiovascular Science, University of Liverpool; School of Primary Care, Population Sciences and Medical Education, University of Southampton; Nutrition and Metabolism, Human Development and Health, Faculty of Medicine, University of Southampton; Southampton National Institute for Health and Care Research, Biomedical Research Centre, University Hospital Southampton

**Keywords:** Heart failure, fibrosis, liver cirrhosis, non-alcoholic fatty liver disease, alcohol

## Abstract

**Background:** Aside from liver related complications, non-alcoholic fatty liver disease (NAFLD) and alcohol-related liver disease (ArLD) are associated with an increased risk of cardiovascular disease (CVD). Liver fibrosis, determined via histology and non-invasive serum fibrosis markers, is associated with cardiovascular events. The association between liver fibrosis and heart failure, and the relationship between *PNPLA3* rs738409 and *TM6SF2* rs58542926 and heart failure is of particular interest, given the known associations of these genetic polymorphisms with increased risk of liver fibrosis and decreased risk of coronary artery disease.

**Methods:** Using data from the UK Biobank (UKBB), we examined the relationship between liver fibrosis, determined using non-invasive markers (NAFLD fibrosis score, Fibrosis-4 (FIB-4) and AST to platelet ratio index (APRI score)) and hospitalization or death from heart failure in 413,860 people. Participants were followed up prospectively via electronic linkage to hospital and death records. Cox-regression estimated the hazard ratios (HR) for death or admission with heart failure. The effects of *PNPLA3* and *TM6SF2* on the association between liver fibrosis and incident heart failure were estimated in an analysis stratified by genotype and by testing for an interaction between genotype and liver fibrosis using a likelihood ratio test.

**Results:** 12,527 incident cases of heart failure occurred over a median of 10.7 years. Liver fibrosis, determined by single or combination non-invasive tests, was associated with an increased risk of hospitalization or death from heart failure; multivariable adjusted high risk NFS score HR 1.59 [1.45-1.76], p<0.0001, FIB-4 HR 1.69 [1.55-1.84], p<0.0001, APRI HR 1.85 [1.56-2.19], p<0.0001, combined fibrosis scores HR 1.90 [1.44-2.49], p<0.0001). These associations persisted for people with NAFLD or harmful alcohol consumption. Polymorphisms linked to liver fibrosis (*PNPLA3* rs738409 GG and *TM6SF2* rs58542926 TT) further amplified the positive association between non-invasive liver fibrosis markers and heart failure. A statistically significant interaction was found between *PNPLA3* rs738409, FIB-4, APRI score and heart failure.

**Conclusion:** Liver fibrosis, determined via non-invasive tests, is associated with an increased risk of hospitalization/death from heart failure in a general population cohort with mixed etiologies of chronic liver disease, including individuals with NAFLD and harmful alcohol consumption. Genetic polymorphisms associated with increased risk of liver fibrosis further increased the risk of heart failure. These findings have important mechanistic, clinical, and public health implications.

## BACKGROUND

Liver disease is a major public health problem globally.^1^ In addition to adverse liver related outcomes (hepatic failure, transplant, liver cancer and death), liver disease is a significant risk factor for adverse cardiovascular outcomes.^2^ An increasing prevalence of obesity and type 2 diabetes (diabesity) has resulted in the emergence of non-alcoholic fatty liver disease (NAFLD) as the leading cause of chronic liver disease. Morbidity and mortality associated with NAFLD is projected to increase exponentially between now and 2030.^3^ NAFLD is a multisystem disease that increases risk of a myriad of other diseases beyond the liver, i.e. significant extra-hepatic complications.^4^ Indeed cardiovascular disease (CVD) is the leading cause of mortality for people with NAFLD,^5^ independent of shared cardiometabolic risk factors.^4,6^ Alcohol-related liver disease (ArLD) is the leading cause of cirrhosis worldwide.^1^ Similarly higher alcohol consumption is associated with an elevated risk of CVD with a roughly linear association with increased risk of stroke, coronary disease (excluding myocardial infarction), heart failure and hypertensive heart disease.^7^ Due to the synergistic effect of metabolic risk factors and alcohol consumption, Europe and North America are currently at high risk of increased liver disease cases in the next few decades.^1^ A potential link between NAFLD and heart failure is a rapidly proliferating research area and an association has been confirmed in several studies.^6,8–10^ A recent meta-analysis of 11 longitudinal cohort studies, including multi-national aggregate data on 11,242,231 middle-aged individuals with 97,716 cases of incident heart failure over a median of 10 years, confirmed that NAFLD is an independent risk factor for incident heart failure (pooled random-effects hazard ratio 1.50, 95% confidence interval 1.34-1.67, p<0.0001), although the authors were not able to examine the association between liver fibrosis and heart failure.^10^

Liver fibrosis is a key histological stage associated with adverse liver-related clinical outcomes for all etiologies of chronic liver disease.^11,12^ Liver fibrosis is also strongly associated with extra-hepatic complications, in particular cardiovascular disease (CVD).^6,13,14^ This association remains irrespective of whether fibrosis is determined histologically^6^ or with non-invasive markers.^13,14^ Very few studies have examined the relationship between liver fibrosis and heart failure however. This is a highly pertinent question, both from a clinical and pathophysiological perspective, given the rising incidence of liver fibrosis in the population and the high levels of morbidity, mortality and health care costs associated with heart failure.

Multiple gene wide association studies (GWAS) and candidate gene studies have identified an association between single nuclear polymorphisms (SNP) in two specific genes: patatin-like phospholipase domain-containing 3 (*PNPLA3*) (rs738409 c.444 C>G, encoding Ile148Met (I148M)) and transmembrane 6 superfamily member 2 (*TM6SF*2) (rs58542926 c.449 C>T, p.Glu167Lys, E167K)) with more advanced liver disease, particularly fibrosis. Thus, those subjects with the GG homozygous variant for *PNPLA3* and the TT homozygous variant for *TM6SF2* are at the highest risk of liver fibrosis. This association is valid in people with NAFLD,^15–18^ and other etiologies of chronic liver disease,^19^ including ArLD,^20–22^ and chronic hepatitis C.^23^ Genetic analysis can be used to identify patients at high risk of liver fibrosis without the inherent issue of metabolic/environmental risk factors confounding any association between liver fibrosis and extra-hepatic complications. Considering the paradox whereby the SNPs rs738409 (*PNPLA3)* and rs58542926 (*TM6SF2*) are associated with liver fibrosis, and yet confer protection against CVD,^24^ the relationship between these genetic variants and incident heart failure is of great interest. These data would help establish whether any causal relationship (or not) exists between liver and cardiac fibrosis and would influence clinical practice.

Using data from the UK Biobank (UKBB), we aimed to examine the relationship between liver fibrosis, determined using both non-invasive markers of liver fibrosis and genetic markers (rs738409 and rs58542926 SNPs), and incident heart failure. We also aimed to test whether there was any modifying influence or interaction, between these genetic markers and the liver fibrosis scores affecting the risk of incident heart failure, in a general population cohort.

## METHODS

### Study population

The UKBB is a longitudinal, prospective study and an unparalleled national health resource aimed at improving and preventing chronic disease through the identification of genetic and behavioral determinants of health in middle-aged and older adults.^25^ Over 500,000 individuals, aged 40-69 years, identified from National Health Service registers agreed to participate and were recruited between 2006 to 2010. At a baseline assessment visit, participants completed a touch-screen questionnaire about their sociodemographic, lifestyle and medical history. Self-reported doctor diagnosed medical conditions and medication history were verified and coded by nurses during a face-to-face interview.^26^ The volunteers underwent a physical examination including anthropometric measurements and provided non-fasting blood and urine samples. Genome-wide genotyping was performed on 488,377 participants using the UK Biobank Axiom Array. UKBB participants gave their consent to be continually followed-up through linkage to electronic health records, including records from the Office for National Statistics and the Registrar General’s Office (death records and cancer registers) in addition to hospital records, held by the Department of Health’s Hospital Episode Statistics and the Scottish Morbidity Records (http://content.digital.nhs.uk/services). At the time of analysis, mortality data were available up to January 2023. Ethical approval was granted by the Northwest Multi-Centre Research Ethics Committee.

### Inclusion and exclusion criteria

All UK Biobank participants were initially included. Exclusions were made for individuals with a baseline ICD code or self-reported UKBB code for heart failure (**Supplementary Methods**), and those for whom we did not have sufficient data to calculate non-invasive markers of fibrosis, including the fibrosis-4 (FIB-4) score,^27^ NAFLD fibrosis score (NFS),^28^ or aspartate transaminase to platelet ratio index (APRI) (**Box 1**).^29^

##### Box 1. Non-invasive fibrosis markers and their calculation

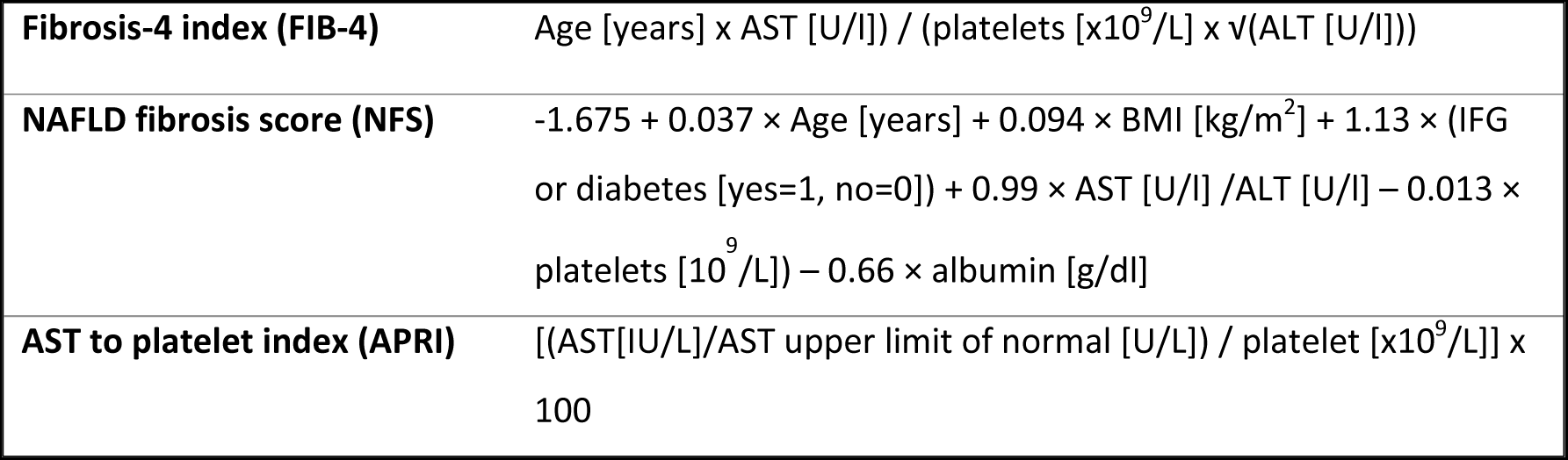

### Ascertainment of exposure

Advanced fibrosis was defined as FIB-4 score > 2.67, NFS > 0.676 or APRI score ≥ 1.0. Advanced fibrosis was excluded if there was a FIB-4 score < 1.3 (< 2.0 if ≥ 65 years), NFS < −1.455 (<0.12 if ≥ 65 years) or APRI < 1.0. Participants not falling into these groups were placed in an indeterminate fibrosis group.

### Primary outcome

The primary outcome for this study was incident heart failure, defined as an ICD-9 (428.0-428.9) or ICD-10 (I50.0-I50.9) code for a new diagnosis of heart failure (i.e., a hospital admission), or heart failure given as the primary, and or secondary cause of death on the death certificate. We investigated a combination of non-fatal and fatal events given the low overall number of heart failure related deaths. Where a participant may have had multiple admissions with heart failure, the first event was included.

### Statistical analysis

Baseline characteristics are presented as percentages, and continuous data are presented using the median and interquartile range (IQR). Univariate and multivariable Cox proportional hazards models were used to calculate the association of elevated non-invasive markers of liver fibrosis on heart failure to determine hazard ratios (HR), 95% confidence intervals (CIs) and statistical significance. Non-cases were censored at the date of loss to follow-up, date of death or the end of follow-up. Data on confounding factors was gathered from the baseline assessment and pre-selected factors identified to be clinically relevant from the literature were entered into multivariable regression models. **Model 1** was adjusted for age, sex, ethnicity and Townsend deprivation index (an address-based metric of socioeconomic status based on car ownership, home ownership, employment and over-crowding).^30^ **Model 2** was additionally adjusted for alcohol (weekly grams; continuous) and smoking status (never, previous, current). **Model 3** was adjusted for the above factors, in addition to metabolic factors including: type 2 diabetes, baseline waist circumference, hypertension, and dyslipidemia. **Model 4** was additionally adjusted for a prior history of acute coronary syndrome, valvular disease, cardiomyopathy and arrhythmias (identified by the participant, or coded for in hospital records). Full definitions are provided in the **Supplementary Methods.** The linearity of the effect of each continuous variable in the adjusted models (i.e., weekly alcohol grams, age, waist circumference, deprivation) was determined using univariate Cox hazard regression with penalized splines. Where a Wald-type test using the nonlinear coefficient estimates indicated significant non-linearity, splines were used in further analyses. The validity of the proportional hazards assumption for each variable was determined by examining correlations between scaled Schoenfeld residuals and time. The statistical package used was R. The Strengthening the Reporting of Observational Studies in Epidemiology (STROBE) guidelines were followed in reporting this study.^31^

### Sensitivity analyses

We performed two sensitivity analyses; the first excluding individuals who were either former, or harmful drinkers (women drinking over 280 grams per week, or men drinking > 400 grams per week), and the second excluding people with baseline cardiovascular disease, including heart failure, acute coronary syndrome, valve disease, cardiomyopathy, arrhythmias.

### Subgroup analyses

We performed the same analysis in participants with a baseline diagnosis of NAFLD, defined as having an ICD-9 (571.8) or ICD-10 (K75.8, K76.0) code indicating NAFLD, or a hepatic steatosis index (HSI) score > 30,^32^ in the absence of another etiology of chronic liver disease; and those with harmful alcohol consumption defined as drinking more than 280g of alcohol per week (women), or over 400g of alcohol per week (men), or having an ICD code for alcohol related liver disease, or and ICD code for alcohol dependency (see **Supplementary Methods** for definitions). In addition, we analyzed the relationship between non-invasive markers of liver fibrosis and heart failure in people living with or without obesity and those living with or without type 2 diabetes (see **Supplementary Methods** for definitions).

### Gene effect analysis

In total 488,377 individuals were genotyped for up to 812,428 variants using DNA extracted from blood samples on either the UKBB Axiom array (438,427 participants) or the UK BiLEVE Axiom array (49,950 participants).^33^ We investigated the effect of the two SNPs of interest: rs738409 (*PNPLA3)* (chromosome 22, location 44324727) and rs58542926 (*TM6SF2*) (chromosome 19, location 19268740) that are known to be associated with liver fibrosis. We assessed the association between liver fibrosis determined via non-invasive tests and hospitalization, or death, from heart failure stratified by these genotypes. The interaction between non-invasive serum fibrosis scores and genotype in relation to heart failure risk was evaluated using a likelihood ratio test. This test determined whether the inclusion of an interaction term in a Cox model estimating the association between fibrosis score and heart failure significantly improved the model fit.

## RESULTS

### Study population and baseline demographics

All 502,506 participants within the UKBB were included. Following the exclusion of individuals with a diagnosis of heart failure at baseline, or insufficient data to calculate the NFS, FIB-4 or APRI score, the final study population consisted of 413,860 participants (**Figure 1**). Baseline demographics are shown in **Table 1**. The baseline characteristics of individuals excluded did not differ significantly to the overall UKBB population (**Supplementary Table 1).** In total 1.3% (n=5400), 2.2% (n=9226) and 0.5% (n=2169) of people in the UKBB had evidence of liver fibrosis according to the NFS, FIB-4 and APRI scores respectively.

**Figure 1.**
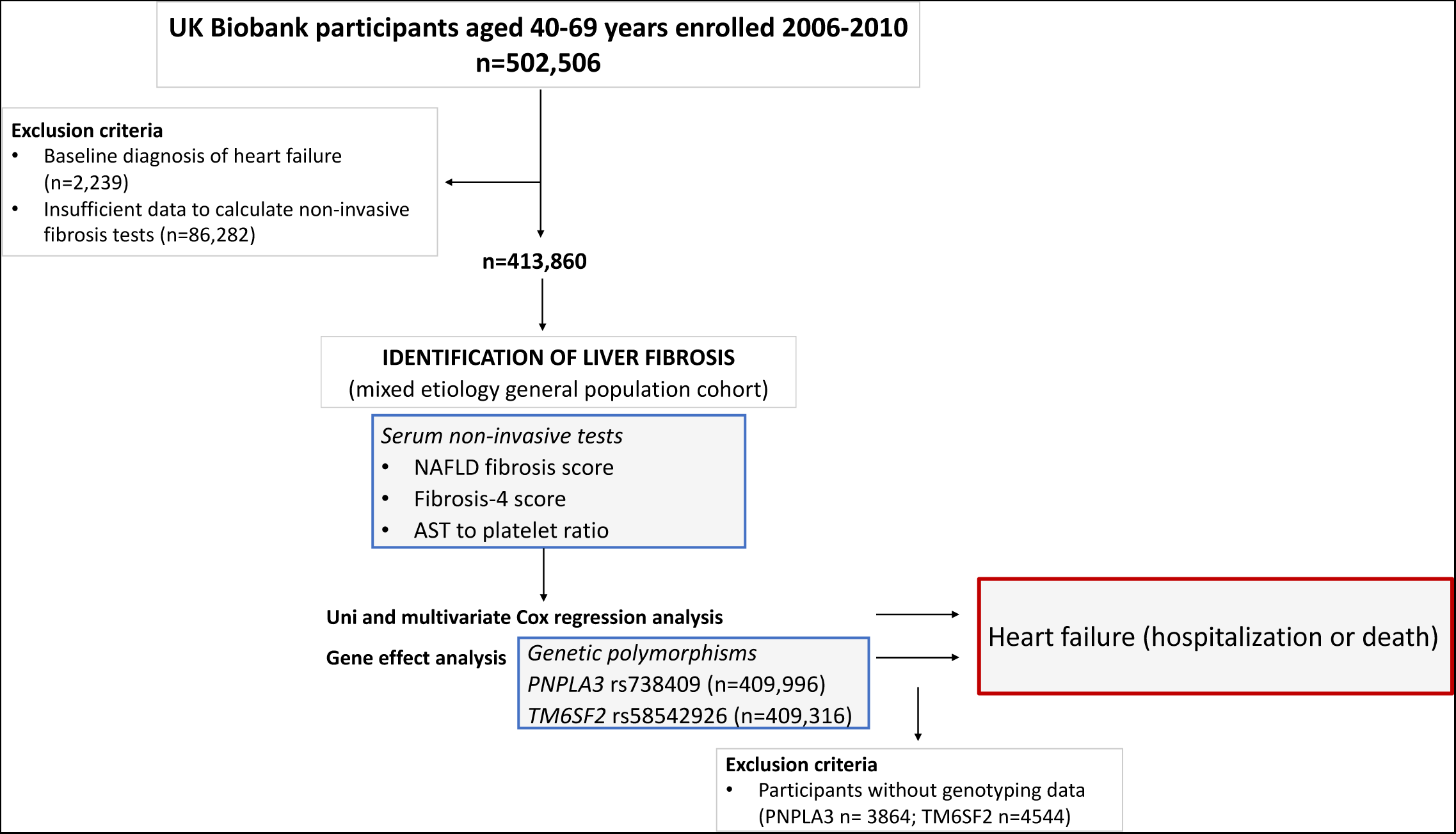
Study flow chart

**Table 1.** Baseline demographics of individuals with and without advanced liver fibrosis according to non-invasive tests (n=413,860)

|  | NAFLD fibrosis score |  |  | Fibrosis-4 score |  |  | AST platelet ratio index |  |
| --- | --- | --- | --- | --- | --- | --- | --- | --- |
|  | Low risk | Intermediate risk | High risk | Low risk | Intermediate risk | High risk | Low risk | High risk |
| **N, (%)** | 327,270 (79.1) | 81,190 (19.6) | 5400 (1.3) | 271,951 (65.7) | 132,683 (32.1) | 9,226 (2.2) | 411,691 (99.5) | 2,169 (0.5) |
| **Median age, years (IQR)** | 57 (15) | 59 (8) | 63 (8) | 56 (16) | 60 (8) | 64 (7) | 58 (13) | 58 (12) |
| **Male (%)** | 44.5 | 51.9 | 57.9 | 42.6 | 52.1 | 65.5 | 46.0 | 67.8 |
| **Ethnicity (%)** |  |  |  |  |  |  |  |  |
| White | 94.8 | 92.4 | 89.9 | 94.3 | 94.4 | 93.3 | 94.3 | 92.4 |
| Non white | 4.8 | 7.1 | 9.3 | 5.4 | 5.2 | 6.1 | 5.3 | 6.8 |
| **Townsend deprivation index** |  |  |  |  |  |  |  |  |
| Median score | -2.22 | -1.92 | -1.19 | -2.12 | -2.24 | -1.99 | -2.16 | -1.43 |
| **Alcohol** |  |  |  |  |  |  |  |  |
| Gram data available (%) | 83.8 | 83.7 | 81.3 | 82.4 | 86.2 | 87.2 | 83.8 | 84.8 |
| Median alcohol grams per week (IQR) | 130 (192) | 120 (212) | 80 (221) | 122 (194) | 132 (206) | 142 (256) | 126 (196) | 201 (364) |
| Non-drinkers (abstainers & former) (%) | 7.4 | 9.3 | 15.9 | 8.1 | 7.5 | 9.4 | 7.9 | 9.1 |
| Low-moderate risk drinkers (%) | 44.4 | 46.6 | 48.7 | 46.4 | 42.3 | 38.8 | 44.9 | 33.0 |
| High risk drinkers (%) | 48.0 | 43.8 | 35.0 | 45.4 | 50.0 | 51.4 | 46.9 | 57.3 |
| **Diabetes status** |  |  |  |  |  |  |  |  |
| T2 diabetes (%) | 2.5 | 15.6 | 58.2 | 5.9 | 5.2 | 10.2 | 5.7 | 17.2 |
| Median HbA1c mmol/mol | 50.9 | 50.2 | 49.3 | 50.8 | 49.3 | 49.2 | 50.3 | 52.8 |
| **Overweight/obesity** |  |  |  |  |  |  |  |  |
| Median BMI, kg/m<sup>2</sup> (IQR) | 26.2 (5) | 29.1 (7) | 34.2 (11) | 27 (6) | 26.2 (6) | 26.7 (6) | 26.7 (6) | 28.4 (7) |
| BMI < 25 kg/m<sup>2</sup> (%) | 37.0 | 18.7 | 8.4 | 30.9 | 37.4 | 34.5 | 33.1 | 24.3 |
| BMI 25-30 kg/m<sup>2</sup> (%) | 44.1 | 38.2 | 20.2 | 43.1 | 42.0 | 40.6 | 42.7 | 38.3 |
| BMI > 30 kg/m<sup>2</sup> (%) | 18.8 | 43.1 | 71.4 | 26.1 | 20.6 | 24.9 | 24.2 | 37.5 |
| Median WC, cm (IQR) | 88 (18) | 96 (20) | 109 (23) | 90 (18) | 89 (18.2) | 92 (19) | 90 (19) | 96.2 (20) |
| WC men > 102 cm, women > 88 cm (%) | 25.8 | 47.1 | 74.1 | 32.9 | 26.0 | 31.2 | 30.6 | 44.0 |
| **Lipids** |  |  |  |  |  |  |  |  |
| Dyslipidemia (%) | 53.2 | 60.9 | 79.3 | 56.6 | 51.6 | 58.2 | 55.0 | 64.9 |
| Median cholesterol (mg/dL) | 5.7 | 5.4 | 4.6 | 5.7 | 5.6 | 5.3 | 5.7 | 5.3 |
| Median LDL (mmol/L) | 3.6 | 3.4 | 2.8 | 3.6 | 3.5 | 3.2 | 3.5 | 3.2 |
| Median HDL (mmol/L) | 1.4 | 1.3 | 1.2 | 1.4 | 1.4 | 1.4 | 1.4 | 1.3 |
| Median TG (mmol/L) | 1.5 | 1.5 | 1.7 | 1.5 | 1.4 | 1.4 | 1.5 | 1.6 |
| **Hypertension (%)** | 27.7 | 39.2 | 67.7 | 29.4 | 31.6 | 45.3 | 30.4 | 46.4 |
| **Smoking (%)** |  |  |  |  |  |  |  |  |
| Never smoked | 55.1 | 52.4 | 45.5 | 54.5 | 54.7 | 49.1 | 54.5 | 44.2 |
| Previous smoker | 33.8 | 37.0 | 44.0 | 33.7 | 36.0 | 40.6 | 34.5 | 39.1 |
| Current smoker | 10.6 | 10.0 | 9.6 | 11.3 | 8.8 | 9.7 | 10.5 | 16.0 |
| **Biochemistry (Median)** |  |  |  |  |  |  |  |  |
| ALT (IU/L) | 20 | 19 | 18 | 20 | 20 | 22 | 20 | 68 |
| AST (IU/L) | 24 | 25 | 26 | 23 | 27 | 33 | 24 | 84 |
| GGT (IU/L) | 26 | 27 | 32 | 27 | 26 | 32 | 26 | 95 |
| Platelets (10<sup>6</sup>/L) | 260 | 205 | 180 | 269 | 212 | 153 | 248 | 155 |
| Albumin (g/L) | 45 | 44 | 43 | 45 | 45 | 45 | 45 | 45 |
| **Baseline cardiovascular events (%)** |  |  |  |  |  |  |  |  |
| Acute coronary syndrome (%) | 2.0 | 3.8 | 9.4 | 2.1 | 3.0 | 6.2 | 2.5 | 5.3 |
| Valvular disease (%) | 0.9 | 1.2 | 2.3 | 0.9 | 1.2 | 2.5 | 1.0 | 1.8 |
| Cardiomyopathy (%) | 0.1 | 0.2 | 0.4 | 0.1 | 0.1 | 0.2 | 0.1 | 0.3 |
| Arrhythmias (%) | 1.6 | 2.3 | 5.0 | 1.5 | 2.2 | 4.9 | 1.8 | 3.1 |

NAFLD, Non-alcoholic fatty liver disease; AST, aspartate transaminase; IQR, interquartile range; HbA1c, glycated hemoglobin; BMI, body mass index; WC, waist circumference; LDL, low density lipoprotein cholesterol; HDL, high density lipoprotein cholesterol; TG, triglycerides; ALT, alanine transaminase; AST, aspartate transaminase; GGT gamma glutamyl transferase
**Low-moderate risk drinkers:** < 140g/week women, < 210g/week men (where frequency only data is available include people drinking < 3x/week)
**High risk drinkers:** > 140g/week women, > 210g/week men (where frequency only data is available include people drinking ≥ 3x/week)
**NAFLD fibrosis score:** low risk < -1.455 (< 0.12 if ≥ 65 yrs); intermediate risk - 1.455 – 0.676 (0.12-0.676 if ≥ 65 yrs); high risk > 0.676
**Fibrosis-4 score:** low risk < 1.3 (<2.0 if ≥ 65 yrs); intermediate risk 1.3-2.67 (2.0-2.67 if ≥ 65 yrs); high risk > 2.67
**AST platelet ratio index:** low risk < 1.0; high risk ≥ 1.0

We attempted to determine an etiology of liver disease for all participants with raised non-invasive markers (**Table 2, Supplementary Table 2**). Frequencies of NAFLD were 56.2%, 31.3% and 30.0%, and frequencies of ArLD were 12.9%, 18.0% and 32.0% for people with a raised NFS score, FIB-4 score and APRI score respectively.

**Table 2.** Etiology of liver disease for participants with advanced fibrosis according to non-invasive tests.

|  | **NAFLD fibrosis score > 0.676, n (%)** | **Fibrosis-4 > 2.67, n (%)** | **AST platelet ratio index ≥ 1.0, n (%)** |
| --- | --- | --- | --- |
| Non-alcoholic fatty liver disease | 2950 (56.2) | 2820 (31.3) | 619 (30.0) |
| Alcohol related liver disease | 680 (12.9) | 1620 (18.0) | 661 (32.0) |
| Viral hepatitis | 39 (0.7) | 87 (1.0) | 63 (3.1) |
| Autoimmune and biliary liver disease | 32 (0.6) | 67 (0.7) | 32 (1.6) |
| Hemochromatosis | 19 (0.4) | 31 (0.3) | 10 (0.5) |
| Other (vascular, drug-induced, inherited liver disease) | 2 (0.04) | 6 (0.1) | 3 (0.1) |
| No clearly identifiable cause of liver disease | 1531 (29.1) | 4374 (48.6) | 676 (32.8) |

NAFLD, Non-alcoholic fatty liver disease; AST, aspartate transaminase
**NAFLD:** HSI > 30 or ICD code for NAFLD (excluding people drinking > 140 grams per week (women) or > 210 grams per week (men), or UKBB or ICD code for alcohol excess, or a non-NAFLD causes of liver disease)
**ArLD:** People drinking at harmful levels (> 280 grams per week women, > 400 grams per week men), or ICD code for alcohol related liver disease, or alcohol dependency
**Viral hepatitis:** UKBB or ICD code for viral hepatitis
**Autoimmune and biliary liver disease:** UKBB or ICD code for autoimmune hepatitis, primary biliary cholangitis, or primary sclerosing cholangitis
**Hemochromatosis:** UKBB or ICD code for haemochromatosis
**Other:** UKBB or ICD code for vascular, drug-induced or inherited liver disease
**No identifiable cause:** Not meeting criteria for any of the above

### Association between non-invasive fibrosis markers and incident heart failure

The median follow-up period for combined fatal and non-fatal incident heart failure was 10.7 years. In total 12,527 cases of hospitalization or death from heart failure were recorded (96 of these were deaths) (**Table 3**). In total, 1926 (15.4%) of heart failure events occurred 12 months following an ICD code for acute coronary syndrome. Only 53 (0.4%) of people who developed heart failure also had a code for alcoholic cardiomyopathy. Elevated levels of non-invasive markers of liver fibrosis at baseline were associated with higher event rates of incident heart failure (**Table 3**).

**Table 3.** Association of the non-invasive tests of liver fibrosis with hospitalization or death from heart failure in a general population cohort.

|  | Risk fibrosis | Participants | Events (fatal) | Median follow up (months) | Event rate | Univariate HR (95% CI) | Model 1 HR (95% CI) | Model 2 HR (95% CI) | Model 3 HR (95% CI) | Model 4 HR (95% CI) |
| --- | --- | --- | --- | --- | --- | --- | --- | --- | --- | --- |
| **NAFLD fibrosis score** | Low | 327,270 | 8248 (55) | 123.8 | 2.19 | 1.00 Ref. | 1.00 Ref. | 1.00 Ref. | 1.00 Ref. | 1.00 Ref. |
|  | Intermediate | 81,190 | 3498 (27) | 128.4 | 0.90 | 1.76 (1.70-1.83) **** | 1.77 (1.7-1.85) **** | 1.77 (1.69-1.85) **** | 1.33 (1.26-1.4) **** | 1.30 (1.24-1.37) **** |
|  | High | 5400 | 781 (14) | 122.4 | 0.21 | 6.25 (5.81-6.73) **** | 3.61 (3.35-3.89) **** | 3.69 (3.39-4.01) **** | 1.72 (1.56-1.9) **** | 1.59 (1.47-1.78) **** |
| **Fibrosis-4 score** | Low | 271,951 | 7514 (59) | 126.8 | 1.95 | 1.00 Ref. | 1.00 Ref. | 1.00 Ref. | 1.00 Ref. | 1.00 Ref. |
|  | Intermediate | 132,683 | 4292 (29) | 129.4 | 1.09 | 1.24 (1.19-1.29) **** | 1.07 (1.03-1.12) *** | 1.13 (1.08-1.18) **** | 1.33 (1.26-1.4) **** | 1.17 (1.12-1.23) **** |
|  | High | 9,226 | 721 (8) | 120.9 | 0.20 | 3.23 (3.00-3.49) **** | 1.66 (1.54-1.8) **** | 1.82 (1.67-1.98) **** | 1.72 (1.56-1.9) **** | 1.69 (1.55-1.84) **** |
| **AST platelet ratio index** | Low | 411,691 | 12366 (94) | 126.1 | 3.22 | 1.00 Ref. | 1.00 Ref. | 1.00 Ref. | 1.00 Ref. | 1.00 Ref. |
|  | High | 2,169 | 161 (2) | 129.1 | 0.04 | 2.68 (2.29-3.13) **** | 2.28 (1.95-2.66) p **** | 2.38 (2.00-2.82) **** | 1.88 (1.58-2.23) **** | 1.85 (1.56-2.19) **** |

NAFLD, Non-alcoholic fatty liver disease; AST, aspartate transaminase; HR, hazard ratio; CI, confidence interval
\* p<0.05, \*\* p<0.01, \*\*\* p<0.001, \*\*\*\* p <0.0001
**NAFLD fibrosis score:** low risk < -1.455 (< 0.12 if ≥ 65 yrs); intermediate risk - 1.455 – 0.676 (0.12-0.676 if ≥ 65 yrs); high risk > 0.676
**Fibrosis-4 score:** low risk < 1.3 (<2.0 if ≥ 65 yrs); intermediate risk 1.3-2.67 (2.0-2.67 if ≥ 65 yrs); high risk > 2.67
**AST platelet ratio index:** low risk < 1.0; high risk ≥ 1.0
**Multivariable model 1:** Adjusted for age, sex, deprivation, ethnicity
**Multivariable model 2:** Adjusted for age, sex, deprivation, ethnicity, alcohol (continuous), smoking
**Multivariable model 3:** Adjusted for age, sex, deprivation, ethnicity, alcohol (continuous), smoking, diabetes, waist circumference, hypertension, dyslipidemia
**Multivariable model 4:** Adjusted for age, sex, deprivation, ethnicity, alcohol (continuous), smoking, diabetes, waist circumference, hypertension, dyslipidemia baseline acute coronary syndrome, valve disease, cardiomyopathy

*Univariate analysis.* Univariate analysis of baseline factors associated with heart failure are shown in **Supplementary Table 3**. Univariate analysis revealed that high NFS (HR 6.25 [5.81-6.73], p<0.0001), FIB-4 (HR 3.23 [3.00-3.49], p<0.0001) and APRI (HR 2.68 [2.29-3.13], p<0.0001) scores were associated with increased risk of incident heart failure (**Table 3**).

*Multivariable analysis.* Following multivariable adjustment for demographics, smoking, alcohol intake, metabolic risk factors and baseline cardiovascular disease, the NFS (HR 1.59 [1.47-1.78], p<0.0001), FIB-4 (HR 1.69 [1.55-1.84], p<0.0001) and APRI scores (HR 1.85 [1.56-2.19], p<0.0001) remained significantly associated with the development of heart failure (**Table 3**, **Figure 2**). A ‘dose response’ relationship by severity was observed for the NFS and FIB-4 scores.

**Figure 2.**
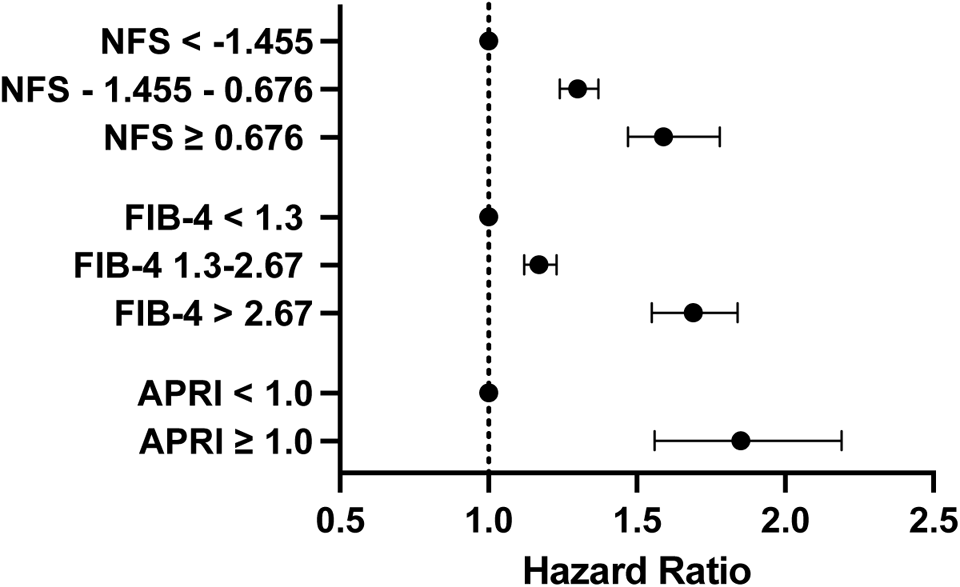
Hazard ratios and 95% confidence intervals for the association of non-invasive markers of liver fibrosis and hospitalization or death due to heart failure following full multivariable adjustment in a general population cohort

*Associations with combinations of more than one high liver fibrosis score.* Where liver fibrosis was defined using combinations of non-invasive tests, people who scored ‘high’ on more than one fibrosis marker were at higher risk of developing heart failure (**Supplementary Table 4, Figure 3**).

**Figure 3.**
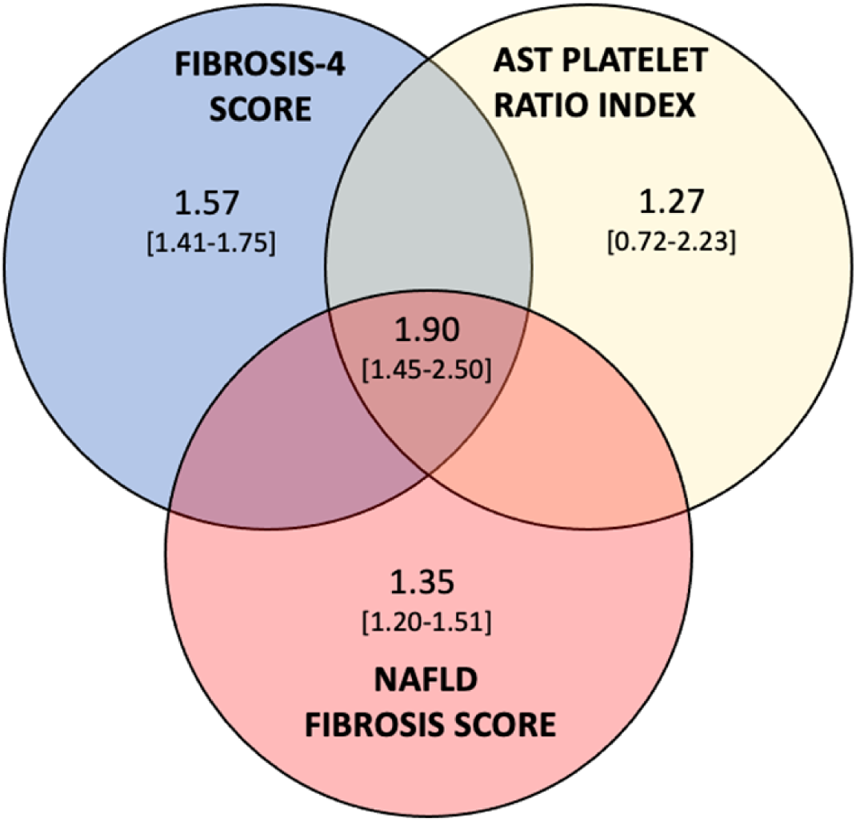
Hazard ratios and 95% confidence intervals for the association of combined non-invasive markers of liver fibrosis and hospitalization or death due to heart failure following full multivariable adjustment in a general population cohort

### Subgroup analysis

*NAFLD and harmful alcohol consumption.* We repeated this analysis in individuals whom we determined were likely to have a baseline diagnosis of NAFLD (n=185,002) and those with evidence of current, or previous harmful alcohol consumption (n=49,156) (see **Supplementary Table 5** for baseline demographics). For NAFLD, a high NFS (HR 1.75 [1.54-1.99], p<0.0001) and FIB-4 score (HR 1.34 [1.14-1.57], p<0.001) were associated with incident heart failure following full multivariable adjustment (**Supplementary Table 6**). For people with harmful alcohol consumption all three fibrosis scores were associated with heart failure after full multivariable adjustment (NFS HR 1.98 [1.59-2.48], p<0.0001; FIB-4 HR 2.08 [1.74-2.49], p<0.0001; APRI HR 2.45 [1.90-3.16], p<0.0001) (**Supplementary Table 7**).

*Type 2 diabetes and obesity status.* All three non-invasive fibrosis scores remained positively associated with heart failure death, or hospitalization, for people with and without obesity (**Supplementary Table 8**). For people with type 2 diabetes, only the NFS score remained associated with heart failure after full multivariable adjustment **(Supplementary Table 9**).

### Sensitivity analyses

We performed a sensitivity analysis excluding individuals who were either former drinkers, or drinking > 280g/week (women), or > 400g/week (men) (**Supplementary Table 10**), and people with baseline cardiovascular disease (**Supplementary Table 11**). For both analyses, all three non-invasive markers of fibrosis continued to be significantly associated with an elevated risk of hospitalization or death from heart failure.

### Genetic effect analysis

*TM6SF2 and PNPLA3 SNPs.* Within our study cohort for *PNPLA3,* 19,576 (4.8%) expressed the homozygous variant GG, 138,229 (33.7%) the heterozygous variant GC and 252,191 (61.5%) CC. For *TM6SF2*, 350,578 (85.6%) carried the polymorphisms CC, 56,473 (13.8%) TC and 2265 (0.6%) TT.

In univariate analysis, there was a trend towards an increase in the risk of incident heart failure for people with evidence of liver fibrosis determined via a non-invasive test for people with *PNPLA3* GG (high risk allele for liver fibrosis) homozygosity compared to those with the heterozygous state, or *PNPLA3* CC homozygosity (**Table 4**). Similarly, there was a trend towards amplified risk for *TM6SF2* heterozygosity compared to *TM6SF2* CC (low risk allele for liver fibrosis) for people with liver fibrosis determined via a high risk FIB-4 or APRI score at baseline (**Table 4**).

**Table 4.** Association of non-invasive markers of liver fibrosis with hospitalization or death from heart failure stratified according to *PNPLA3* and *TM6SF2* polymorphisms in a general population cohort (univariate analysis)

|  |  |  | NAFLD fibrosis score |  |  | Fibrosis-4 score |  |  | AST platelet ratio index |  |
| --- | --- | --- | --- | --- | --- | --- | --- | --- | --- | --- |
|  |  |  | Low | Intermediate | High | Low | Intermediate | High | Low | High |
| ***PNPLA3***<br>(rs738409) | CC | No. cases HF | 5150 | 2159 | 466 | 4790 | 2583 | 402 | 7697 | 78 |
|  |  | HR (95% CI) | 1.00 Ref. | 1.72 (1.64-1.81) **** | 6.20 (5.64- 6.82) **** | 1.00 Ref. | 1.20 (1.14- 1.25) **** | 2.99 (2.70- 3.31) **** | 1.00 Ref. | 2.68 (2.15- 3.35) **** |
|  | CG<br>GC | No. cases HF | 2690 | 1143 | 269 | 2386 | 1455 | 261 | 4046 | 56 |
|  |  | HR (95% CI) | 0.96 (0.92- 1.01) | 1.74 (1.63- 1.85) **** | 5.90 (5.21- 6.67) **** | 0.94 (0.89- 0.98) ** | 1.21 (1.14- 1.29) **** | 3.28 (2.89- 3.71) **** | 0.97 (0.94- 1.01) | 2.20 (1.69- 2.87) **** |
|  | GG | No. cases HF | 342 | 169 | 41 | 283 | 216 | 53 | 526 | 26 |
|  |  | HR (95% CI) | 0.89 (0.80- 0.99) * | 1.89 (1.63- 2.21) **** | 7.67 (5.64-10.43) **** | 0.84 (0.75- 0.95) ** | 1.26 (1.10- 1.44) *** | 4.02 (3.06- 5.26) **** | 0.94 (0.86- 1.02) | 4.70 (3.20- 6.90) **** |
| ***TM6SF2***<br>(rs58542926) | CC | No. cases HF | 7000 | 2981 | 674 | 6388 | 3672 | 595 | 10,530 | 125 |
|  |  | HR (95% CI) | 1.00 Ref. | 1.75 (1.68- 1.83) **** | 6.28 (5.80- 6.80) **** | 1.00 Ref. | 1.24 (1.19- 1.29) **** | 3.17 (2.91- 3.45) **** | 1.00 Ref. | 2.54 (2.13- 3.03) **** |
|  | CT<br>TC | No. cases HF | 1132 | 476 | 95 | 1025 | 561 | 117 | 1671 | 32 |
|  |  | HR (95% CI) | 1.01 (0.95- 1.07) | 1.93 (1.76- 2.12) **** | 6.16 (5.03- 7.54) **** | 1.00 (0.94- 1.07) | 1.26 (1.16- 1.38) **** | 3.73 (3.10- 4.48) **** | 1.01 (0.96- 1.06) | 3.52 (2.49- 4.98) **** |
|  | TT | No. cases HF | 33 | 16 | 6 | 35 | 18 | 2 | 53 | 2 |
|  |  | HR (95% CI) | 0.76 (0.54- 1.08) | 1.51 (0.93- 2.47) | 6.59 (2.96-14.67) **** | 0.89 (0.64- 1.24) | 1.00 (0.63- 1.59) | 1.17 (0.29- 4.67) | 0.82 (0.63- 1.08) | 2.33 (0.58- 9.33) |

Grey shaded boxes represent risk alleles associated with an increased risk of liver fibrosis
NAFLD, Non-alcoholic fatty liver disease; AST, aspartate transaminase; HR, hazard ratio; CI, confidence interval; HF, heart failure.
\* p<0.05, \*\* p<0.01, \*\*\* p<0.001, \*\*\*\* p <0.0001
**NAFLD fibrosis score:** low risk < -1.455 (< 0.12 if ≥ 65 yrs); intermediate risk - 1.455 – 0.676 (0.12-0.676 if ≥ 65 yrs); high risk > 0.676
**Fibrosis-4 score:** low risk < 1.3 (<2.0 if ≥ 65 yrs); intermediate risk 1.3-2.67 (2.0-2.67 if ≥ 65 yrs); high risk > 2.67
**AST platelet ratio index:** low risk < 1.0; high risk ≥ 1.0

We incorporated interaction terms into our models to examine how *PNPLA3* and *TM6SF2* influence the relationships between fibrosis scores and heart failure risk. The interactions involving *PNPLA3* rs738409 with both FIB-4 and APRI were statistically significant. No statistically significant interaction was found with *TM6SF2* although the overall number of people with *TM6SF2* TT homozygosity and a heart failure event were very small.

## DISCUSSION

### Summary of the main findings

We have identified that liver fibrosis, determined by non-invasive tests, was associated with an elevated risk of hospitalization or death from heart failure, following full multivariable adjustment in a prospective general population cohort, unselected for any type, or risk of liver disease. This association persists for people identified to have either NAFLD or harmful alcohol consumption, and for people living with or without obesity or type 2 diabetes. Polymorphisms known to identify people at risk of liver fibrosis (*PNPLA3* rs738409 GG and *TM6SF2* rs58542926 TT) were associated with a further increase in risk of incident heart failure despite these variants having been shown to be associated with a decreased risk of coronary artery disease.^24^ This was particularly apparent for fibrosis *PNPLA3* rs738409 GG where a statistically significant interaction was found between this SNP and a high liver fibrosis score, to further amplify the risk of incident heart failure.

### Comparison to the literature

While few studies have directly explored the relationship between liver fibrosis and incident heart failure, data from the literature supports our findings. In the Multi-Ethnic Study of Atherosclerosis (cross-sectional, six US centres), using cardiac and liver T1-mapping magnetic resonance imaging, liver fibrosis was found to be associated with a with history of heart failure and with preclinical measures of abnormal cardiac structure and function, including left ventricular hypertrophy and myocardial fibrosis.^34^ Using UKBB data, we recently reported that in a population of people with chronic kidney disease who also had NAFLD, elevated non-invasive markers of liver fibrosis are associated with heart failure.^35^ Simon *et al* demonstrated in a prospective study of > 10,000 Swedish adults, followed up for a median of 13 years, that there was an increased risk of major adverse cardiovascular events in people with NAFLD.^6^ In this study, 153 people developed congestive heart failure amongst the 1554 people with NAFLD who had non-cirrhotic fibrosis and there was an adjusted hazard ratio of 2.04 [1.66-2.51]. This study benefited from liver histological data to diagnose liver fibrosis but the overall numbers of individuals experiencing heart failure were relatively small and residual unmeasured confounding may have existed. Data from the Korean National Health Insurance identified that liver fibrosis, defined using the BARD score, in people with background NAFLD, determined using the fatty liver index, was associated with increased incident and hospitalized heart failure.^36^ To our knowledge, no previous studies have examined the relationship between liver fibrosis and heart failure for patients in a general population, nor have any previous studies tested whether genetic polymorphisms known to identify people at risk of liver fibrosis (*PNPLA3* rs738409 GG and *TM6SF2* rs58542926 TT) modify risk of incident heart failure. This is important as these variants have previously been shown to be associated with a decreased risk of coronary artery disease.^24^

We have demonstrated that liver fibrosis, determined via non-invasive tests, in people with an alcohol-related etiology is a risk factor for heart failure. As far as we are aware, ours is also the first study to identify this association. Chronic high levels of alcohol consumption can lead to a dilated cardiomyopathy (alcoholic cardiomyopathy) characterized by cardiomegaly, reduced contractility, and disruption of myofibrillary architecture. However, only 0.4% of people with a heart failure outcome in this study had an ICD code for alcoholic cardiomyopathy and therefore there may be shared mechanistic pathways across liver disease etiologies. Furthermore, a proportion of individuals in this UKBB cohort will have heart failure related to cirrhotic cardiomyopathy. This is a distinct syndrome which results from the release of proinflammatory signals and circulatory abnormalities inherent to patients with cirrhosis, in particular those with decompensated disease. We anticipate that this would only account for a minor number of cases however, given that this is a general population study recruiting healthy volunteers.

We provide novel insight by exploring the association of non-invasive fibrosis markers with heart failure stratified according to genetic polymorphisms associated with liver fibrosis. *PNPLA3* rs738409 GG is known to be associated with liver fibrosis in people with NAFLD.^15,16^ Candidate gene studies from Europe have shown that the E167K variant of *TM6SF2* rs58542926 is associated with an almost two-fold increase in the risk of advanced fibrosis independent of age, diabetes, obesity, or *PNPLA3* genotype.^17,18^ In addition to NAFLD, rs738409 for *PNPLA3* and rs58542926 for *TM6SF2* have been linked to ArLD, in particular alcohol-related cirrhosis,^20–22^ more advanced liver disease for patients with chronic hepatitis C,^23^ and progressive fibrosis in patients with different etiologies of chronic liver disease.^19,22^ While these genetic variants confer increased risk for people developing more advanced liver disease, they paradoxically confer protection against cardiovascular disease.^24^ An exome-wide association study has shown that the *TM6SF2* TT and *PNPLA3 GG* genotypes are involved in hepatic production of triglyceride-rich lipoproteins, and are associated with increased risk of type 2 diabetes and a lower risk of coronary artery disease.^24^ This protection likely results from decreased circulating levels of very low density lipoprotein (VLDL) due to reduced export of this lipoprotein from the liver, reducing the impact of the atherogenic lipoprotein phenotype that occurs with NAFLD on CVD effects.^37^ Three metabolomic signature subgroups have been identified for NAFLD which align with CVD risk; subtype A which exhibits impaired VLDL-triglyceride secretion is associated with lower risk of CVD.^38^ It might therefore

### Clinical relevance

Understanding the potential mechanistic link(s) between liver fibrosis and heart failure has significant therapeutic relevance and clinical and public health consequences. Heart failure represents a major public health problem with a globally increasing prevalence largely due to a combination of an ageing population with worsening metabolic health from the obesity epidemic; ischemic and hypertensive heart disease represent the leading causes in both men and women.^40^ Patients’ health are increasingly characterised by multimorbidity, further exacerbated by socioeconomic deprivation.^41^ Heart failure is associated with high levels of morbidity and mortality and contributes significantly towards high health care expenditure and frequent hospital admissions.^42^ There is predicted to be a substantial rise in population levels of liver fibrosis driven by NAFLD and alcohol consumption,^1^ with a likely associated impact on cardiovascular disease including heart failure. Early lifestyle intervention (reduced alcohol consumption, improved nutrition, and regular physical activity) and pharmacological treatments (in development), in those identified with liver fibrosis, may prevent progression towards clinically significant cardiac disease including heart failure. NAFLD-specific therapies which can reverse and prevent progression to liver fibrosis (from steatosis/steatohepatitis) are beginning to be realised.^43^ There is however also a need to address the huge burden of undiagnosed chronic liver disease in the community.^44^ Widespread screening of at-risk clinical groups represents a paradigm shift in clinical practice and will strain existing health care systems. Non-invasive serum markers represent a useful first line screening approach, found to be predictive of liver related morbidity in the general population.^45^ There is not yet clear guidance as to whether patients with liver fibrosis should be screened for CVD, despite the acknowledgement that NAFLD contributes towards CVD risk and shares risk factors with CVD.^46–48^ European guidelines for NAFLD advocate screening for CVD although they do not stipulate which assessments should occur,^47^ whereas the American guidelines support screening for metabolic risk factors alone.^46^ Findings from this study provide support for targeted screening for fibrosis in people with risk factors for liver disease and the adoption of a cardiometabolic multimorbidity approach to the management of people with established liver fibrosis focusing on modifiable shared risk factors.

### Strengths and weaknesses

The UKBB is a substantial (half a million participants) prospective multi-modality dataset with nearly 17 years of clinical event follow-up time. Baseline data are available to calculate multiple well validated prediction scores for risk stratification for advanced liver fibrosis, enabling us to examine the incremental risk between levels of exposure (liver fibrosis) and incident heart failure. We were additionally able to adjust for multiple potential confounders and identify incident cases of both fatal and non-fatal heart failure using linked datasets. Finally, using this specific cohort we were able to undertake a genetic association study to further validate this association and investigate the potential modifying influence of SNPs in two genes known to be associated with increased risk of liver fibrosis.

The accurate identification of liver fibrosis is challenging, with even a gold standard liver biopsy assessment subject to sampling and analysis bias.^49^ Even with the limitations of liver biopsy, this would not be feasible, nor ethically acceptable, in the UKBB cohort without a clinical indication. Non-invasive blood-based tests achieve high negative, but low positive predictive values, although test combinations enhance diagnostic accuracy as utilized here. Simple non-invasive tests have been shown however to perform as well as histology in predicting clinical outcomes (a composite endpoint of all-cause mortality and liver related events) in patients with NAFLD.^50^ False positive results may have resulted from non-hepatic conditions for example hematological disorders or infection. Indeed, it was not possible to identify a driver of chronic liver disease in all participants with a raised fibrosis marker. It is possible that participants with a high BMI or diabetes who did not have an HSI > 30 had NAFLD, or people with reported high-risk (but not harmful) levels of alcohol intake, or a high frequency of alcohol intake without weekly gram data had ArLD, however we wanted to be conservative in our definitions to allow accurate subgroup analysis. We were reliant on ICD coding for heart failure and therefore our results fail to capture the large proportion of people with incident heart failure not associated with a hospital admission or death, i.e. less advanced disease. Admission and mortality data identifies the spectrum of heart failure associated with high levels of mortality and health care costs however, and is thus where prevention is most urgently needed.

Stratification of univariate associations of non-invasive markers of liver fibrosis with heart failure according to SNPs linked to liver fibrosis, revealed a trend towards an amplified risk for *PNPLA3* rs738409 GG, however the number of cases of heart failure were small for people in the group at the greatest risk (high risk non-invasive fibrosis marker and *PNPLA3* rs738409 GG) leading to wide confidence intervals. This issue was more pronounced for *TM6SF2* rs58542926 TT, where the number of events was extremely small, however a similar trend can be seen for the heterozygous state compared to *TM6SF2* rs58542926 CC where the event numbers were greater. Analysis of the interaction of *TM6SF2* rs58542926 was also likely underpowered given the low number of people with *TM6SF2* TT homozygosity. We were unable to elucidate the relationship between liver fibrosis and different phenotypes of heart failure (heart failure with either preserved or reduced ejection fraction). While patients with heart failure are at increased risk of congestive hepatopathy and ischemic hepatitis which can influence non-invasive fibrosis marker scores, fibrosis markers were calculated at baseline and individuals with pre-existing heart failure were excluded, in addition to those with underlying CVD, at the study entry point in a further sensitivity analysis which did not alter the overall findings. Sequential data on liver fibrosis markers is only available for a small proportion of UKBB participants therefore we are unable to perform time dependent regression modelling. Finally, being an observational study, we were unable to derive mechanistic insight into the link between liver fibrosis and heart failure, an area of undoubted future research relevant in the quest to identify novel therapeutic targets in the liver-cardiac axis.

## Conclusion

Liver fibrosis, determined via non-invasive tests, is associated with an increased risk of hospitalization or death from heart failure in a general population where liver fibrosis was associated with a mixed etiology of chronic liver disease, including individuals with NAFLD and harmful alcohol consumption. Genetic polymorphisms associated with increased risk of liver fibrosis further increased the risk of heart failure. Stratification of risk of heart failure based on liver fibrosis genotype has also potential future relevance in an era of personalised medicine. These data provide a rationale for examining the feasibility and effectiveness of targeted community screening for undetected liver disease followed by multimorbidity assessment and aggressive cardiovascular risk factor management to prevent the development of heart failure in people with hepatic fibrosis.

## Funding

CDB is supported in part by the Southampton NIHR Biomedical Research Centre (NIHR203319), UK.

## Disclosures

None

## Data Availability

Data is available for download following submission of an applicaiton to the UK Biobank

## Acknowledgements

None

## Notes

### Competing Interest Statement

The authors have declared no competing interest.

### Author Declarations

Ethical approval was granted by the Northwest Multi-Centre Research Ethics Committee.

